# Effectiveness of BNT162b2 XBB vaccine in the US Veterans Affairs Healthcare System

**DOI:** 10.1101/2024.04.05.24305063

**Authors:** Aisling R. Caffrey, Haley J. Appaneal, Vrishali V. Lopes, Laura Puzniak, Evan J. Zasowski, Luis Jodar, Kerry L. LaPlante, John M. McLaughlin

## Abstract

Data evaluating effectiveness of XBB.1.5-adapted vaccines against JN.1-related endpoints are scarce. We performed a nationwide test-negative case-control study within the US Veterans Affairs Healthcare System to estimate vaccine effectiveness (VE) of BNT162b2 XBB.1.5-adapted vaccine compared to not receiving an XBB vaccine of any kind against COVID-19 hospitalization, emergency department or urgent care visits (ED/UC), and outpatient visits. Between September 25, 2023 and January 31, 2024, effectiveness was 24–35% during a period of JN.1 predominance and 50–61% during XBB predominance across all outcomes. VE within 60 days of vaccination during the likely JN.1 period was 32% (95% confidence interval 3-52%) against hospitalization, 41% (23–54%) against ED/UC visits, and 31% (1–52%) against outpatient visits. Corresponding VE during the likely XBB period was 62% (44–74%), 52% (37–63%), and 50% (25–66%) by setting, respectively. These data underscore the importance of strain match to maximize the public health impact of COVID-19 vaccination.

## BACKGROUND

Coronavirus disease 2019 (COVID-19), caused by the SARS-CoV-2 virus, continues to pose significant global health challenges more than four years after its emergence. COVID-19 still causes hundreds of thousands of deaths annually worldwide, and millions of survivors have long-term sequalae following acute COVID-19.^1,2^ Highly effective vaccines targeting SARS-CoV-2 have been key in blunting the public health impact of COVID-19 over the past several years.^3–8^ Variant-adapted versions of the vaccines have been developed to maintain protection against COVID-19 as SARS-CoV-2 continues to evolve.^9–13^ Monovalent mRNA vaccines adapted to target the XBB.1.5 sub-lineage (hereafter referred to as XBB vaccines) were authorized or approved for all individuals ≥6 months of age by the United States Food and Drug Administration on September 11^th^, 2023.^14^ Data from the early portion of the 2023–2024 respiratory season indicated that XBB vaccines have been effective at preventing COVID-19, including severe disease.^15–19^

JN.1, which is antigenically and phylogenetically distinct from XBB sub-lineages, was first detected in late August of 2023 and became the predominant circulating variant in the United States and elsewhere by mid-to-late December 2023.^20^ JN.1 has shown some signs of immune-escape,^21–23^ which in turn could reduce the effectiveness of current XBB vaccines. Data evaluating the effectiveness of XBB vaccines against JN.1 are both scarce and urgently needed to help guide regulators and public health policy about the need for COVID- 19 vaccine strain updates prior to the 2024–2025 respiratory virus season.^17^ We evaluated the effectiveness of the Pfizer-BioNTech 2023–2024 formulation, which was a monovalent XBB.1.5-containing vaccine (hereafter referred to as BNT162b2 XBB vaccine), across time periods of XBB and JN.1 sub-lineage predominance among adults in a large US nationwide integrated healthcare system.

## METHODS

### Setting and Participants

We conducted a nationwide test-negative case-control study using clinical data from patients of the US Veterans Affairs (VA) Healthcare System. We assessed the effectiveness of the BNT162b2 XBB vaccine among adult patients ≥18 years of age diagnosed with an acute respiratory infection (ARI; see **Supplemental Table 1**) in the hospital, emergency department (ED), urgent care (UC), or outpatient setting (in-person or virtual) between September 25, 2023 and January 31, 2024. To be included, patients had to be tested for SARS-CoV-2 via nucleic acid amplification test (NAAT) or rapid antigen test (RAT) within 14 days prior through 3 days after the ARI encounter. All ARI encounters within a 30-day window were considered a single ARI episode and the encounter at the highest level of care (i.e., hospitalization > ED/UC visit > outpatient visit) was selected for inclusion. Patients were excluded if they (1) did not have at least one visit to the VA Healthcare System in the previous 12 months, (2) had another prior positive SARS-CoV-2 test in the 90 days prior to their ARI episode, (3) received an XBB vaccine other than BNT162b2, (4) received BNT162b2 XBB vaccine within 8 weeks of a prior COVID-19 vaccine dose, (5) received BNT162b2 XBB vaccine within 14 days of their ARI episode, (6) received BNT162b2 XBB vaccine but the date of administration was unknown, or (7) received a COVID-19 antiviral (nirmatrelvir/ritonavir, remdesivir, or molnupiravir) within 30 days of their ARI episode. Patients could contribute more than one ARI episode to the study if the episodes were more than 30 days apart.

### Outcomes

Three mutually exclusive ARI episode outcome categories were assessed: (1) hospital admission, (2) ED or UC visit (without subsequent hospital admission), and (3) outpatient visits (without a subsequent ED/UC visit or hospital admission). Within each ARI outcome category, cases were those with a positive SARS-CoV-2 NAAT or RAT result, and controls were those who tested negative.

### Exposure

The exposure of interest was receipt of the BNT162b2 XBB vaccine at least 14 days before the ARI episode. Those who had not received an XBB vaccine of any kind were considered unexposed, regardless of prior COVID-19 vaccination history and including those who were not previously vaccinated. Vaccine exposure was evaluated using the VA integrated electronic health record, which captures data across all healthcare settings, including all vaccines administered.^24^ COVID-19 vaccines were offered free of charge to all Veterans enrolled in VA healthcare based on Centers for Disease Control and Prevention (CDC) recommendations during the study period.^25^

### Statistical Analyses

The primary analysis estimated vaccine effectiveness (VE) of BNT162b2 XBB vaccine against hospitalization, ED/UC visits, and outpatient visits separately compared to not receiving an XBB vaccine of any kind. Effectiveness was estimated overall (September 25, 2023 through January 31, 2024) and during three time periods based on the predominantly circulating SARS-CoV-2 strains which were determined using GISAID variant tracking data for the United States^26^: (1) likely XBB period defined as September 25, 2023 through November 30, 2023; (2) XBB and JN.1 co-circulation period defined as December 1, 2023 through December 31, 2023; and (3) likely JN.1 period defined as January 1, 2024 through January 31, 2024. XBB and JN.1 sub- lineages accounted for >90% and >80% of sequenced SARS-CoV-2 strains in the United States during our likely XBB and likely JN.1 periods, respectively.^26^

Separate multivariable logistic regression models were used to compare the odds of receiving a BNT162b2 XBB vaccine between SARS-CoV-2 positive cases and test-negative controls within each ARI outcome category (i.e., hospitalization, ED/UC visit, outpatient visit) and variant period while adjusting for potential confounding variables. Adjusted odds ratios (OR) and corresponding 95% Wald confidence intervals (CI) were constructed. VE was calculated as 1 minus the corresponding adjusted OR (and 95% CI), multiplied by 100%. The following variables were selected a priori based on previous literature, and controlled for in each multivariable logistic regression model: calendar week of ARI episode, age (18–64, 65–74, ≥75 years), sex (male or female), race (Black, White, or other race), ethnicity (Hispanic or non-Hispanic), body mass index (BMI) categories (underweight, healthy weight, overweight, obese, missing), Charlson Comorbidity Index (0, 1, 2, 3, ≥ 4), receipt of influenza vaccine during the 2023–2024 season (yes or no), receipt of pneumococcal vaccine in the past 5 years (yes or no), encounters with the VA healthcare system in the year prior (intensive care unit admission, hospital admission, nursing home admission, ED visit, primary care visit; 0 or ≥1 for each), smoking status (current or former smoker or never smoker), immunocompromised (yes or no), and Census region (Northeast, Midwest, South, or West),^12^ and prior documented SARS-CoV-2 infection.

In secondary analyses, we evaluated adjusted VE by time since BNT162b2 XBB vaccination within both the likely XBB and likely JN.1 time periods to simultaneously assess the impact of potential waning of protection and variant predominance. We calculated adjusted VE estimates within 60 and 61–133 days since vaccination for the likely XBB and JN.1 time periods.

Additional stratified VE analyses were also conducted by age group (<65 and ≥65 years), immunocompromised status (yes or no),^27^ obesity status (BMI ≥30 kg/m^2^ and <30 kg/m^2^),^28^ and smoking status (current or former smoker and never smoker). All logistic regression models were carefully checked for assumptions and model fit. All analyses were conducted using SAS (Version 9.2, SAS Institute Inc., Cary, NC, USA).

## RESULTS

This study included 113,174 ARI episodes with corresponding SARS-CoV-2 test results (**Figure 1**), of which 24,206 (21.4%) were hospital admissions, 61,976 (54.8%) were ED/UC visits, and 26,992 (23.9%) were outpatient visits. Median age was 65 years (interquartile range [IQR]: 52–75; **Table 1**). Most ARI episodes occurred among males (98,172; 86.7%) and those who were White (71,345; 63%); 29,699 (26.2%) were Black or African American, 29,083 (25.7%) had a Charlson Comorbidity Index ≥4, and 40,309 (35.6%) were immunocompromised. Overall, 18.1% (20,523/113,174) were SARS-CoV-2 test positive cases and 81.9% (92,651/113,174) were test-negative controls. A total of 6.5% (7,324/113,174) received BNT162b2 XBB vaccine with a median time since receipt of 54 days (range: 15–133; IQR: 35–76). Of cases and controls, 1,019 (5.0%) and 6,305 (6.8%), respectively, had ever received BNT162b2 XBB vaccine. Most (105,850/113,174 [93.5%]) had never received an XBB vaccine of any kind, and 24,747 (21.9%) had never received a COVID- 19 vaccine of any kind. Among those who received BNT162b2 XBB vaccine, median time since receipt of their most recent previous (non-XBB) dose of COVID-19 vaccine was 433 days (IQR: 385–477).

**Figure 1.**
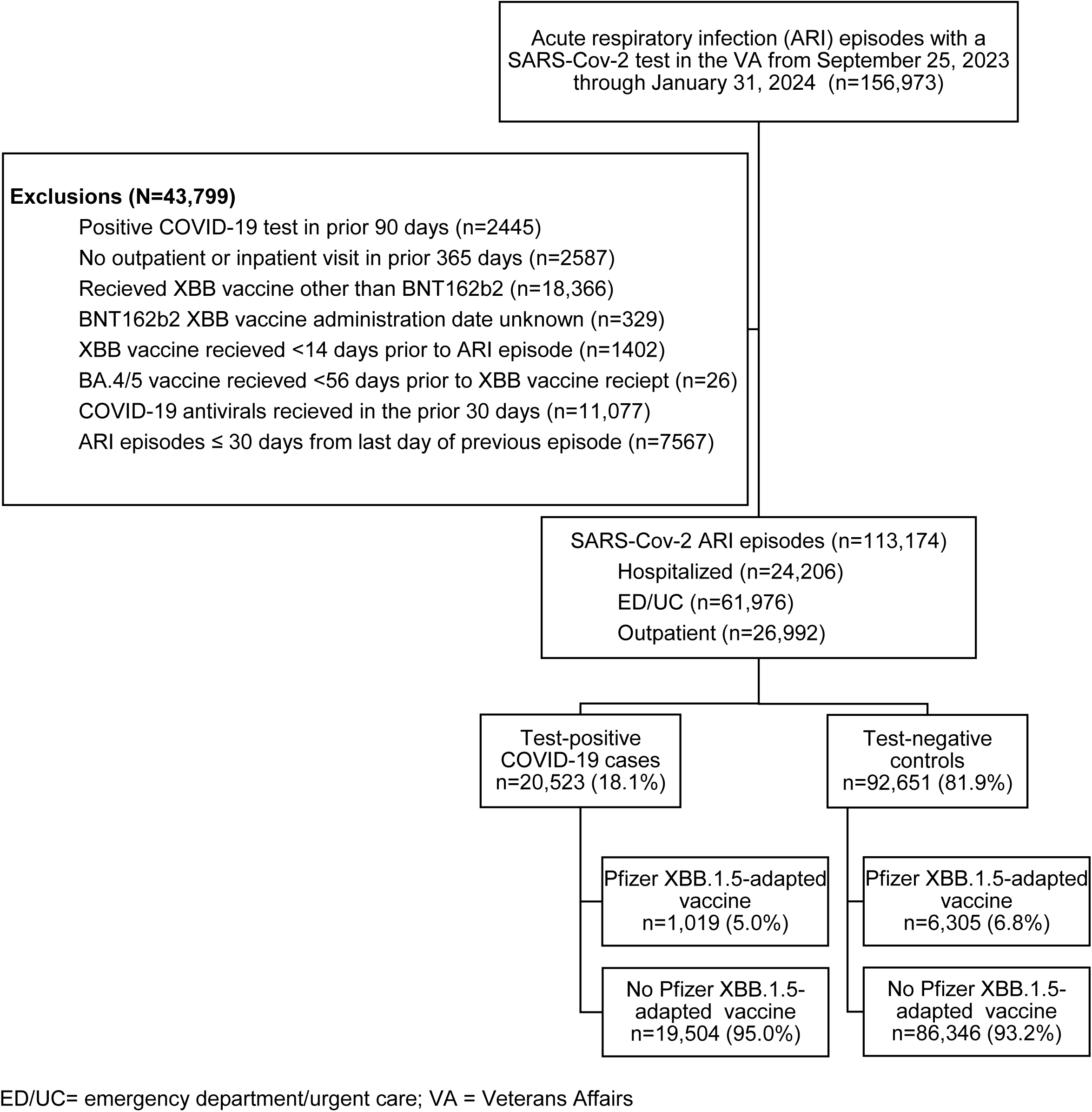
Study selection criteria

**Table 1.**
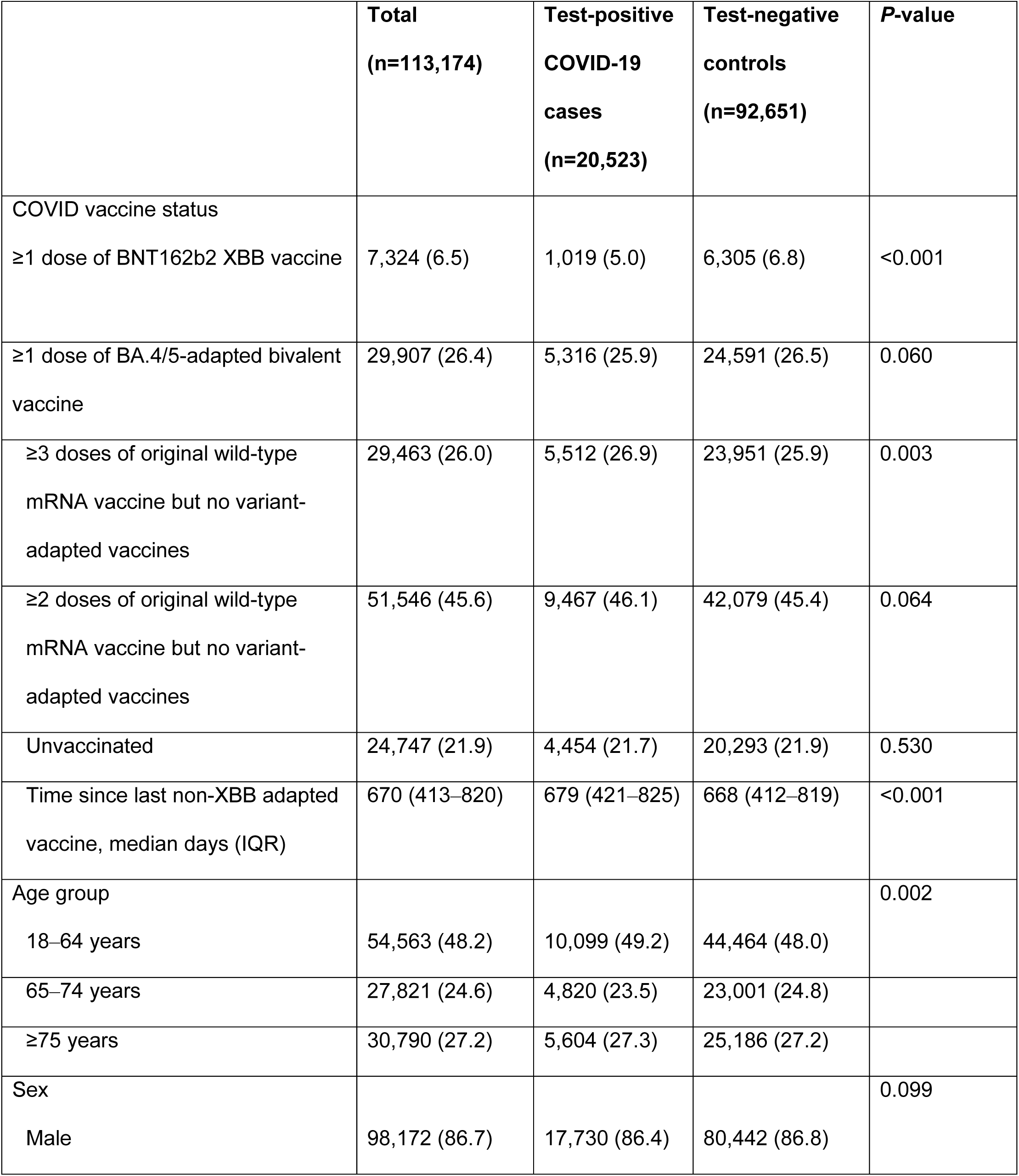
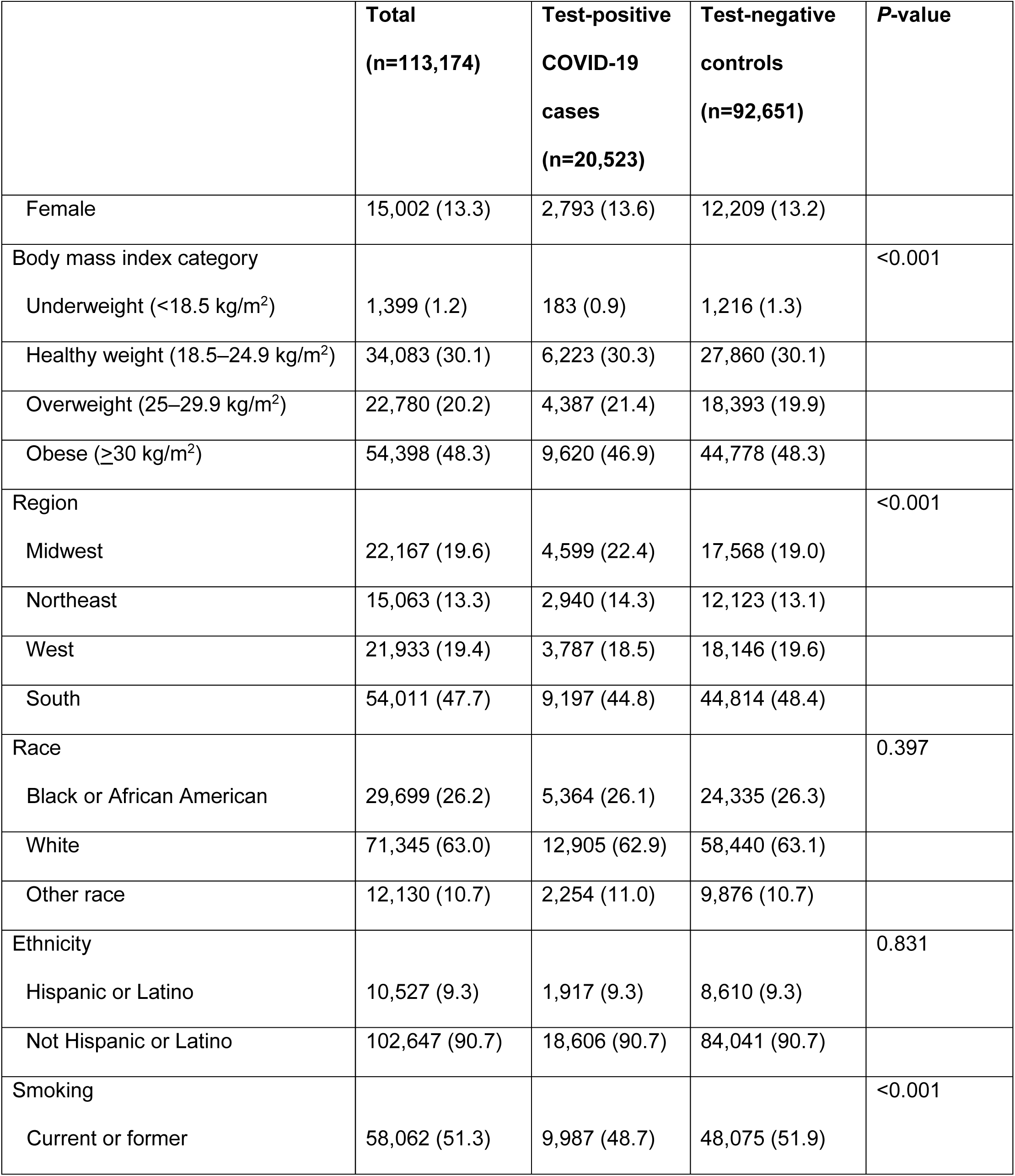
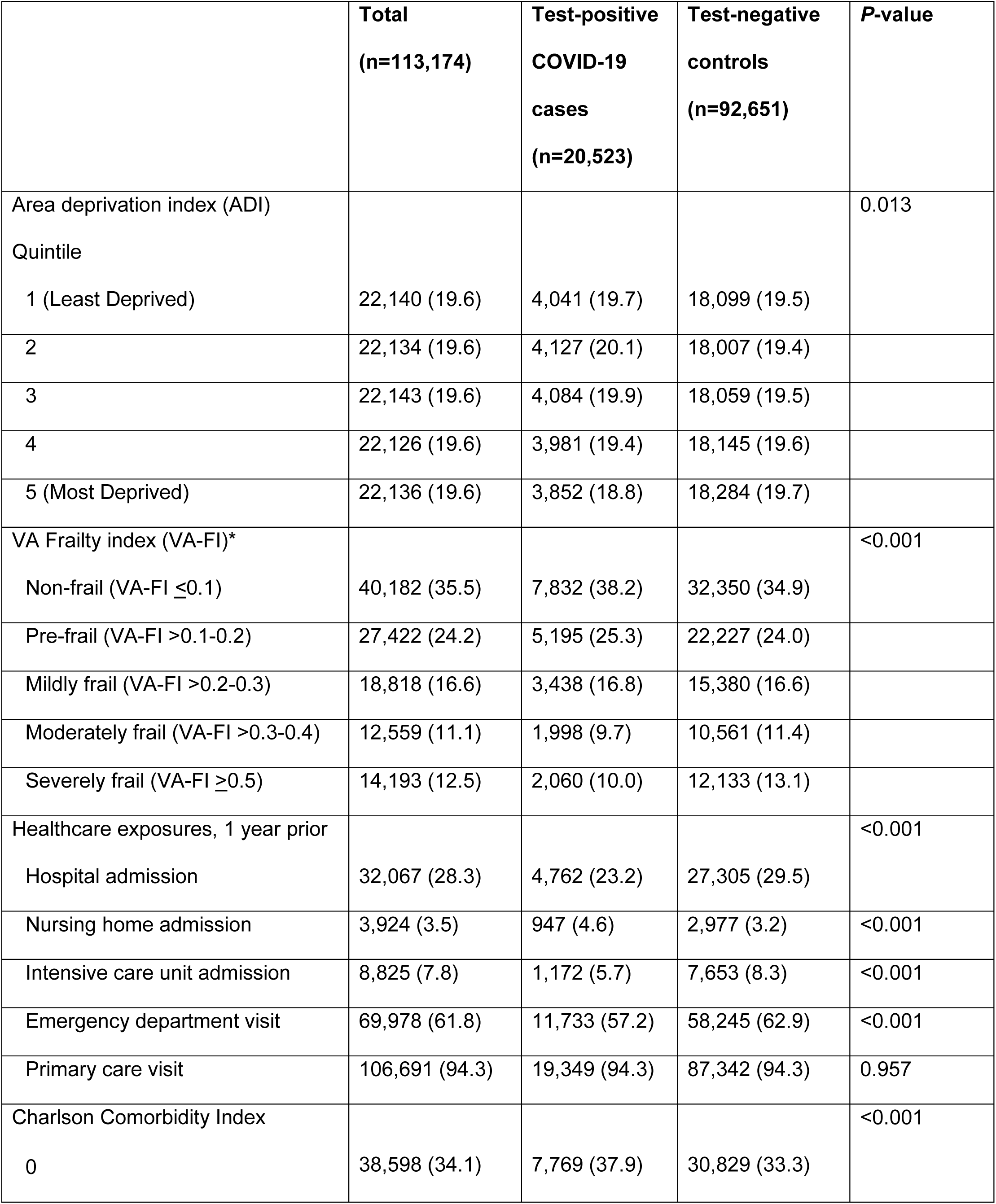
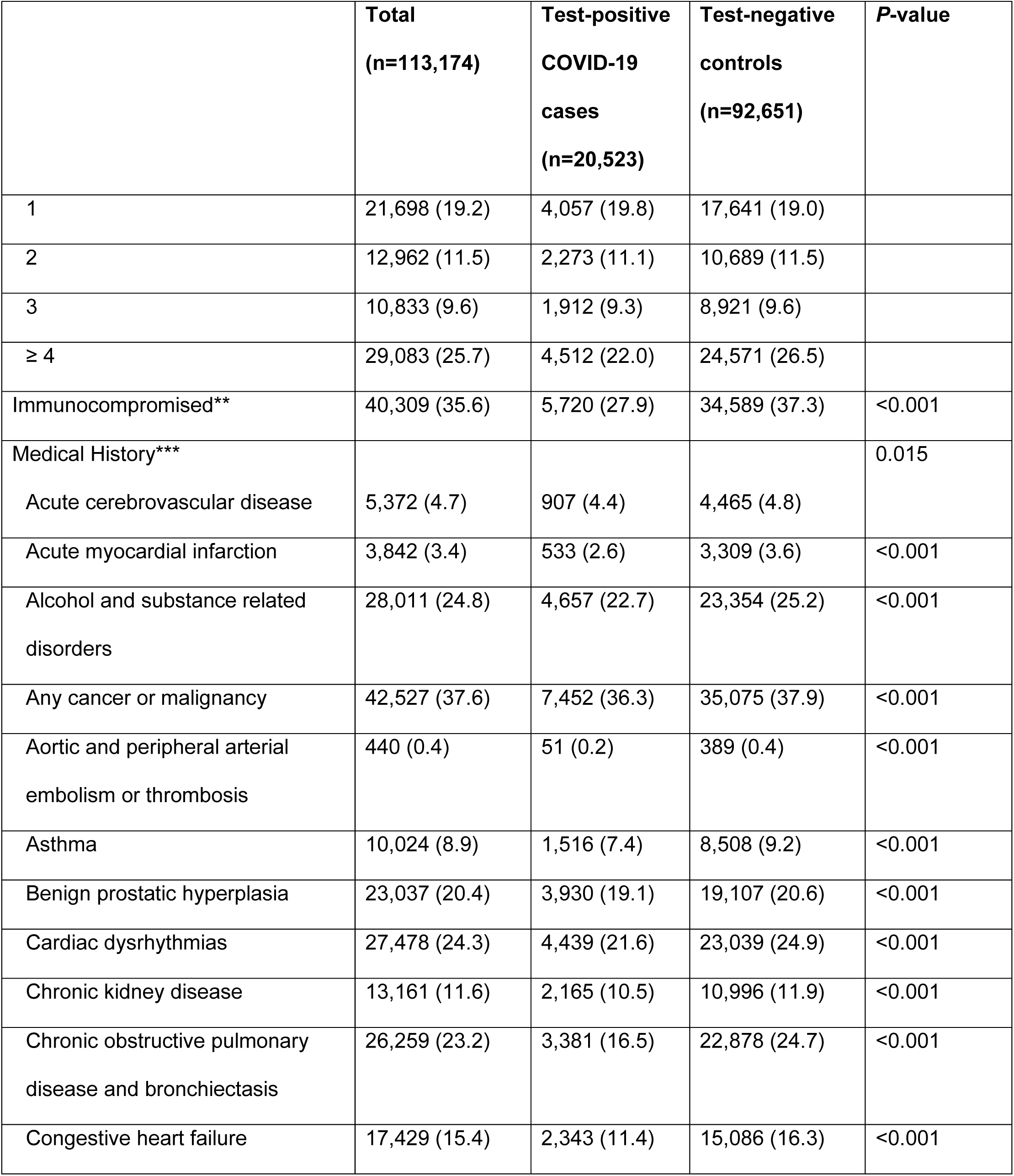
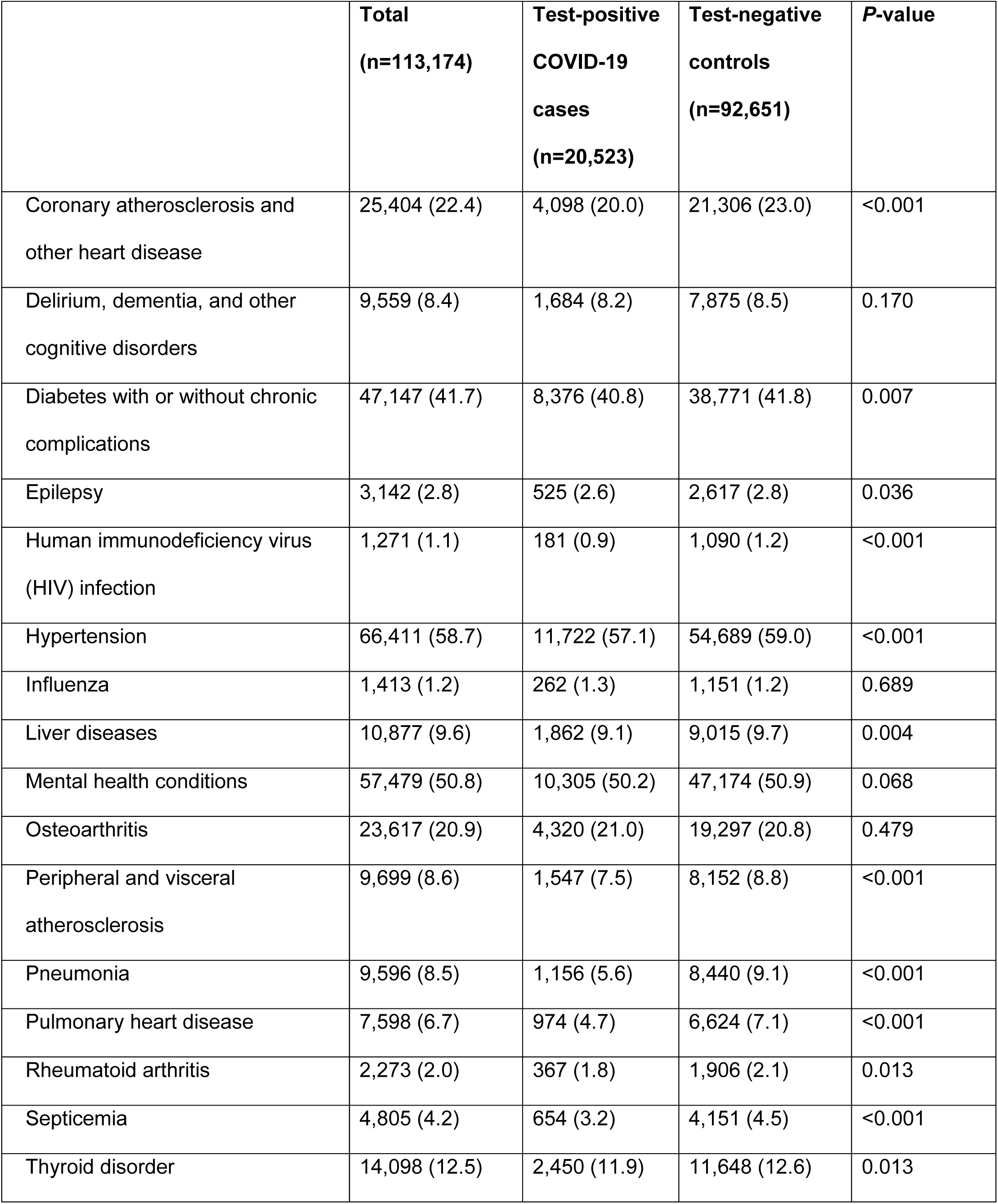
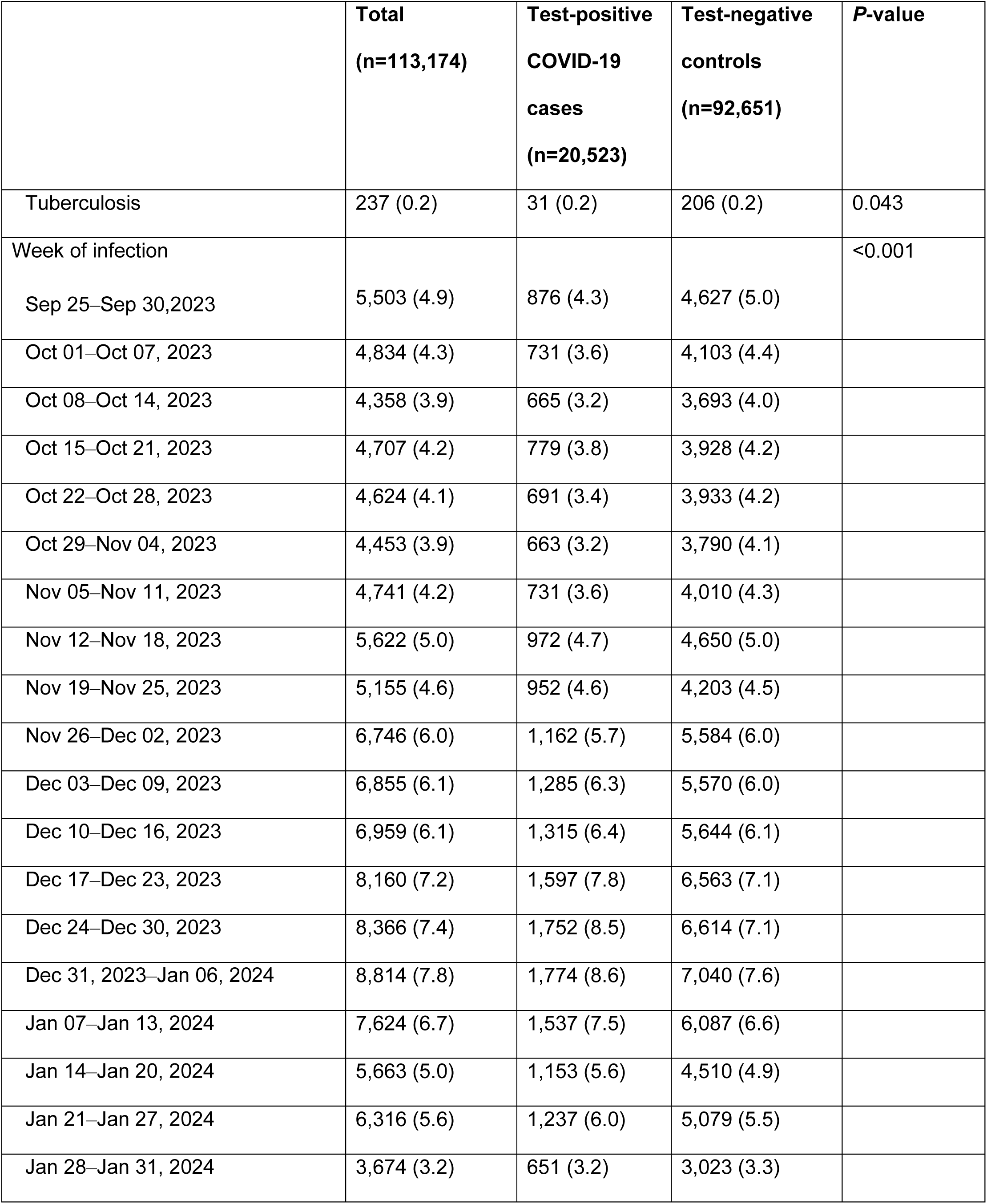
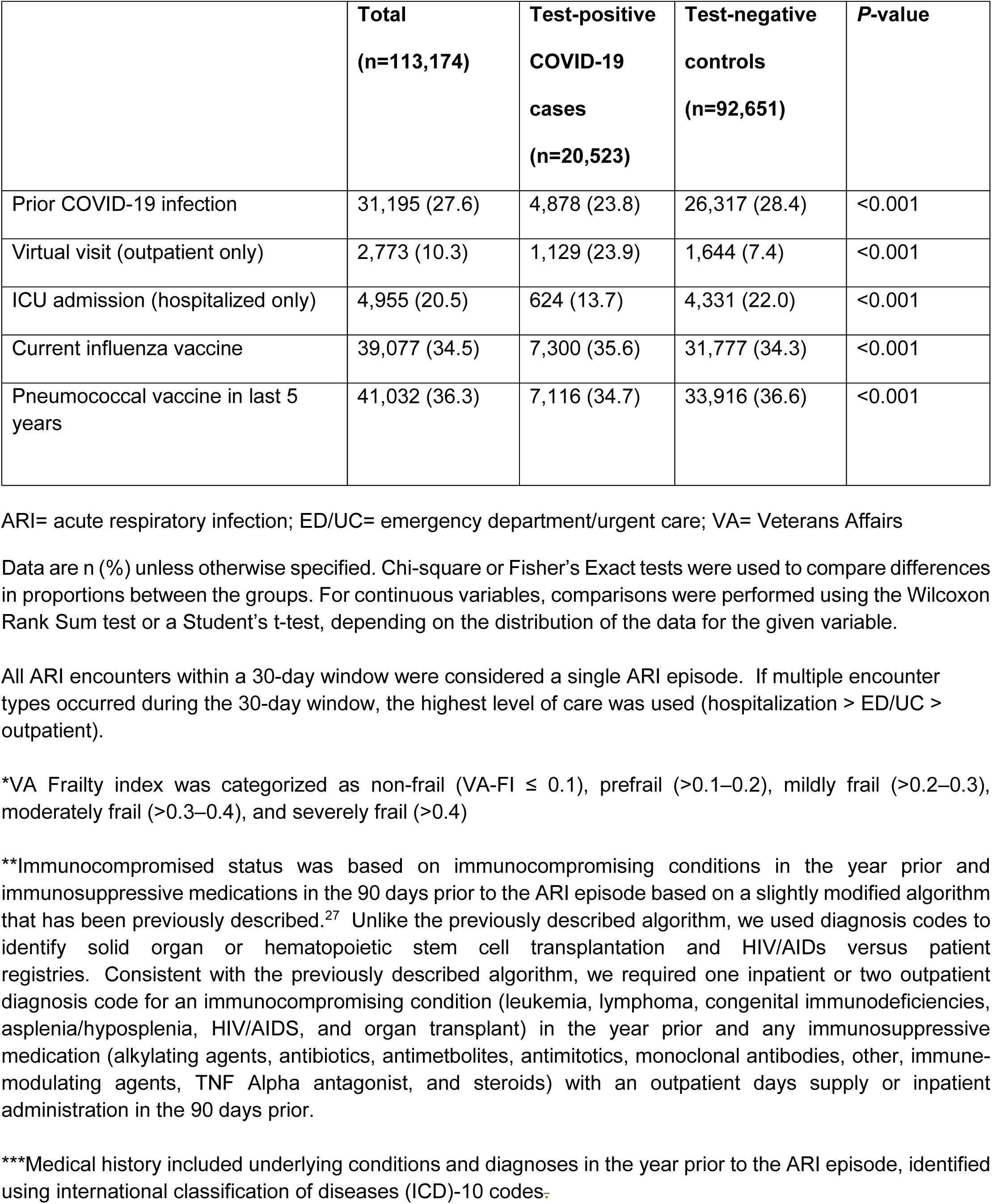
Demographics and clinical characteristics of acute respiratory infection episodes (hospitalization, ED/UC visits, outpatient visits) with SARS-CoV-2 testing by COVID-19 case-control status.

|  | **Total**<br>**(n=113,174)** | **Test-positive**<br>**COVID-19**<br>**cases**<br>**(n=20,523)** | **Test-negative**<br>**controls**<br>**(n=92,651)** | **P-value** |
| --- | --- | --- | --- | --- |
| COVID vaccine status |  |  |  |  |
| ≥1 dose of BNT162b2 XBB vaccine | 7,324 (6.5) | 1,019 (5.0) | 6,305 (6.8) | <0.001 |
| ≥1 dose of BA.4/5-adapted bivalent vaccine | 29,907 (26.4) | 5,316 (25.9) | 24,591 (26.5) | 0.060 |
| ≥3 doses of original wild-type mRNA vaccine but no variant-adapted vaccines | 29,463 (26.0) | 5,512 (26.9) | 23,951 (25.9) | 0.003 |
| ≥2 doses of original wild-type mRNA vaccine but no variant-adapted vaccines | 51,546 (45.6) | 9,467 (46.1) | 42,079 (45.4) | 0.064 |
| Unvaccinated | 24,747 (21.9) | 4,454 (21.7) | 20,293 (21.9) | 0.530 |
| Time since last non-XBB adapted vaccine, median days (IQR) | 670 (413–820) | 679 (421–825) | 668 (412–819) | <0.001 |
| Age group |  |  |  | 0.002 |
| 18–64 years | 54,563 (48.2) | 10,099 (49.2) | 44,464 (48.0) |  |
| 65–74 years | 27,821 (24.6) | 4,820 (23.5) | 23,001 (24.8) |  |
| ≥75 years | 30,790 (27.2) | 5,604 (27.3) | 25,186 (27.2) |  |
| Sex |  |  |  | 0.099 |
| Male | 98,172 (86.7) | 17,730 (86.4) | 80,442 (86.8) |  |
| Female | 15,002 (13.3) | 2,793 (13.6) | 12,209 (13.2) |  |
| Body mass index category |  |  |  | <0.001 |
| Underweight (<18.5 kg/m<sup>2</sup>) | 1,399 (1.2) | 183 (0.9) | 1,216 (1.3) |  |
| Healthy weight (18.5–24.9 kg/m<sup>2</sup>) | 34,083 (30.1) | 6,223 (30.3) | 27,860 (30.1) |  |
| Overweight (25–29.9 kg/m<sup>2</sup>) | 22,780 (20.2) | 4,387 (21.4) | 18,393 (19.9) |  |
| Obese (≥30 kg/m<sup>2</sup>) | 54,398 (48.3) | 9,620 (46.9) | 44,778 (48.3) |  |
| Region |  |  |  | <0.001 |
| Midwest | 22,167 (19.6) | 4,599 (22.4) | 17,568 (19.0) |  |
| Northeast | 15,063 (13.3) | 2,940 (14.3) | 12,123 (13.1) |  |
| West | 21,933 (19.4) | 3,787 (18.5) | 18,146 (19.6) |  |
| South | 54,011 (47.7) | 9,197 (44.8) | 44,814 (48.4) |  |
| Race |  |  |  | 0.397 |
| Black or African American | 29,699 (26.2) | 5,364 (26.1) | 24,335 (26.3) |  |
| White | 71,345 (63.0) | 12,905 (62.9) | 58,440 (63.1) |  |
| Other race | 12,130 (10.7) | 2,254 (11.0) | 9,876 (10.7) |  |
| Ethnicity |  |  |  | 0.831 |
| Hispanic or Latino | 10,527 (9.3) | 1,917 (9.3) | 8,610 (9.3) |  |
| Not Hispanic or Latino | 102,647 (90.7) | 18,606 (90.7) | 84,041 (90.7) |  |
| Smoking |  |  |  | <0.001 |
| Current or former | 58,062 (51.3) | 9,987 (48.7) | 48,075 (51.9) |  |
| Area deprivation index (ADI) |  |  |  | 0.013 |
| Quintile |  |  |  |  |
| 1 (Least Deprived) | 22,140 (19.6) | 4,041 (19.7) | 18,099 (19.5) |  |
| 2 | 22,134 (19.6) | 4,127 (20.1) | 18,007 (19.4) |  |
| 3 | 22,143 (19.6) | 4,084 (19.9) | 18,059 (19.5) |  |
| 4 | 22,126 (19.6) | 3,981 (19.4) | 18,145 (19.6) |  |
| 5 (Most Deprived) | 22,136 (19.6) | 3,852 (18.8) | 18,284 (19.7) |  |
| VA Frailty index (VA-FI)* |  |  |  | <0.001 |
| Non-frail (VA-FI ≤0.1) | 40,182 (35.5) | 7,832 (38.2) | 32,350 (34.9) | |
| Pre-frail (VA-FI >0.1-0.2) | 27,422 (24.2) | 5,195 (25.3) | 22,227 (24.0) |  |
| Mildly frail (VA-FI >0.2-0.3) | 18,818 (16.6) | 3,438 (16.8) | 15,380 (16.6) |  |
| Moderately frail (VA-FI >0.3-0.4) | 12,559 (11.1) | 1,998 (9.7) | 10,561 (11.4) |  |
| Severely frail (VA-FI ≥0.5) | 14,193 (12.5) | 2,060 (10.0) | 12,133 (13.1) | |
| Healthcare exposures, 1 year prior |  |  |  | <0.001 |
| Hospital admission | 32,067 (28.3) | 4,762 (23.2) | 27,305 (29.5) |  |
| Nursing home admission | 3,924 (3.5) | 947 (4.6) | 2,977 (3.2) | <0.001 |
| Intensive care unit admission | 8,825 (7.8) | 1,172 (5.7) | 7,653 (8.3) | <0.001 |
| Emergency department visit | 69,978 (61.8) | 11,733 (57.2) | 58,245 (62.9) | <0.001 |
| Primary care visit | 106,691 (94.3) | 19,349 (94.3) | 87,342 (94.3) | 0.957 |
| Charlson Comorbidity Index |  |  |  | <0.001 |
| 0 | 38,598 (34.1) | 7,769 (37.9) | 30,829 (33.3) |  |
| 1 | 21,698 (19.2) | 4,057 (19.8) | 17,641 (19.0) |  |
| 2 | 12,962 (11.5) | 2,273 (11.1) | 10,689 (11.5) |  |
| 3 | 10,833 (9.6) | 1,912 (9.3) | 8,921 (9.6) |  |
| ≥ 4 | 29,083 (25.7) | 4,512 (22.0) | 24,571 (26.5) |  |
| Immunocompromised** | 40,309 (35.6) | 5,720 (27.9) | 34,589 (37.3) | <0.001 |
| Medical History*** |  |  |  | 0.015 |
| Acute cerebrovascular disease | 5,372 (4.7) | 907 (4.4) | 4,465 (4.8) |  |
| Acute myocardial infarction | 3,842 (3.4) | 533 (2.6) | 3,309 (3.6) | <0.001 |
| Alcohol and substance related disorders | 28,011 (24.8) | 4,657 (22.7) | 23,354 (25.2) | <0.001 |
| Any cancer or malignancy | 42,527 (37.6) | 7,452 (36.3) | 35,075 (37.9) | <0.001 |
| Aortic and peripheral arterial embolism or thrombosis | 440 (0.4) | 51 (0.2) | 389 (0.4) | <0.001 |
| Asthma | 10,024 (8.9) | 1,516 (7.4) | 8,508 (9.2) | <0.001 |
| Benign prostatic hyperplasia | 23,037 (20.4) | 3,930 (19.1) | 19,107 (20.6) | <0.001 |
| Cardiac dysrhythmias | 27,478 (24.3) | 4,439 (21.6) | 23,039 (24.9) | <0.001 |
| Chronic kidney disease | 13,161 (11.6) | 2,165 (10.5) | 10,996 (11.9) | <0.001 |
| Chronic obstructive pulmonary disease and bronchiectasis | 26,259 (23.2) | 3,381 (16.5) | 22,878 (24.7) | <0.001 |
| Congestive heart failure | 17,429 (15.4) | 2,343 (11.4) | 15,086 (16.3) | <0.001 |
| Coronary atherosclerosis and other heart disease | 25,404 (22.4) | 4,098 (20.0) | 21,306 (23.0) | <0.001 |
| Delirium, dementia, and other cognitive disorders | 9,559 (8.4) | 1,684 (8.2) | 7,875 (8.5) | 0.170 |
| Diabetes with or without chronic complications | 47,147 (41.7) | 8,376 (40.8) | 38,771 (41.8) | 0.007 |
| Epilepsy | 3,142 (2.8) | 525 (2.6) | 2,617 (2.8) | 0.036 |
| Human immunodeficiency virus (HIV) infection | 1,271 (1.1) | 181 (0.9) | 1,090 (1.2) | <0.001 |
| Hypertension | 66,411 (58.7) | 11,722 (57.1) | 54,689 (59.0) | <0.001 |
| Influenza | 1,413 (1.2) | 262 (1.3) | 1,151 (1.2) | 0.689 |
| Liver diseases | 10,877 (9.6) | 1,862 (9.1) | 9,015 (9.7) | 0.004 |
| Mental health conditions | 57,479 (50.8) | 10,305 (50.2) | 47,174 (50.9) | 0.068 |
| Osteoarthritis | 23,617 (20.9) | 4,320 (21.0) | 19,297 (20.8) | 0.479 |
| Peripheral and visceral atherosclerosis | 9,699 (8.6) | 1,547 (7.5) | 8,152 (8.8) | <0.001 |
| Pneumonia | 9,596 (8.5) | 1,156 (5.6) | 8,440 (9.1) | <0.001 |
| Pulmonary heart disease | 7,598 (6.7) | 974 (4.7) | 6,624 (7.1) | <0.001 |
| Rheumatoid arthritis | 2,273 (2.0) | 367 (1.8) | 1,906 (2.1) | 0.013 |
| Septicemia | 4,805 (4.2) | 654 (3.2) | 4,151 (4.5) | <0.001 |
| Thyroid disorder | 14,098 (12.5) | 2,450 (11.9) | 11,648 (12.6) | 0.013 |
| Tuberculosis | 237 (0.2) | 31 (0.2) | 206 (0.2) | 0.043 |
| Week of infection |  |  |  | <0.001 |
| Sep 25–Sep 30, 2023 | 5,503 (4.9) | 876 (4.3) | 4,627 (5.0) |  |
| Oct 01–Oct 07, 2023 | 4,834 (4.3) | 731 (3.6) | 4,103 (4.4) |  |
| Oct 08–Oct 14, 2023 | 4,358 (3.9) | 665 (3.2) | 3,693 (4.0) |  |
| Oct 15–Oct 21, 2023 | 4,707 (4.2) | 779 (3.8) | 3,928 (4.2) |  |
| Oct 22–Oct 28, 2023 | 4,624 (4.1) | 691 (3.4) | 3,933 (4.2) |  |
| Oct 29–Nov 04, 2023 | 4,453 (3.9) | 663 (3.2) | 3,790 (4.1) |  |
| Nov 05–Nov 11, 2023 | 4,741 (4.2) | 731 (3.6) | 4,010 (4.3) |  |
| Nov 12–Nov 18, 2023 | 5,622 (5.0) | 972 (4.7) | 4,650 (5.0) |  |
| Nov 19–Nov 25, 2023 | 5,155 (4.6) | 952 (4.6) | 4,203 (4.5) |  |
| Nov 26–Dec 02, 2023 | 6,746 (6.0) | 1,162 (5.7) | 5,584 (6.0) |  |
| Dec 03–Dec 09, 2023 | 6,855 (6.1) | 1,285 (6.3) | 5,570 (6.0) |  |
| Dec 10–Dec 16, 2023 | 6,959 (6.1) | 1,315 (6.4) | 5,644 (6.1) |  |
| Dec 17–Dec 23, 2023 | 8,160 (7.2) | 1,597 (7.8) | 6,563 (7.1) |  |
| Dec 24–Dec 30, 2023 | 8,366 (7.4) | 1,752 (8.5) | 6,614 (7.1) |  |
| Dec 31, 2023–Jan 06, 2024 | 8,814 (7.8) | 1,774 (8.6) | 7,040 (7.6) |  |
| Jan 07–Jan 13, 2024 | 7,624 (6.7) | 1,537 (7.5) | 6,087 (6.6) |  |
| Jan 14–Jan 20, 2024 | 5,663 (5.0) | 1,153 (5.6) | 4,510 (4.9) |  |
| Jan 21–Jan 27, 2024 | 6,316 (5.6) | 1,237 (6.0) | 5,079 (5.5) |  |
| Jan 28–Jan 31, 2024 | 3,674 (3.2) | 651 (3.2) | 3,023 (3.3) |  |
| Prior COVID-19 infection | 31,195 (27.6) | 4,878 (23.8) | 26,317 (28.4) | <0.001 |
| Virtual visit (outpatient only) | 2,773 (10.3) | 1,129 (23.9) | 1,644 (7.4) | <0.001 |
| ICU admission (hospitalized only) | 4,955 (20.5) | 624 (13.7) | 4,331 (22.0) | <0.001 |
| Current influenza vaccine | 39,077 (34.5) | 7,300 (35.6) | 31,777 (34.3) | <0.001 |
| Pneumococcal vaccine in last 5 years | 41,032 (36.3) | 7,116 (34.7) | 33,916 (36.6) | <0.001 |

ARI= acute respiratory infection; ED/UC= emergency department/urgent care; VA= Veterans Affairs
Data are n (%) unless otherwise specified. Chi-square or Fisher's Exact tests were used to compare differences in proportions between the groups. For continuous variables, comparisons were performed using the Wilcoxon Rank Sum test or a Student's t-test, depending on the distribution of the data for the given variable.
All ARI encounters within a 30-day window were considered a single ARI episode. If multiple encounter types occurred during the 30-day window, the highest level of care was used (hospitalization > ED/UC > outpatient).
\*VA Frailty index was categorized as non-frail (VA-FI ≤ 0.1), prefrail (>0.1–0.2), mildly frail (>0.2–0.3), moderately frail (>0.3–0.4), and severely frail (>0.4)
\*\*Immunocompromised status was based on immunocompromising conditions in the year prior and immunosuppressive medications in the 90 days prior to the ARI episode based on a slightly modified algorithm that has been previously described.<sup>27</sup> Unlike the previously described algorithm, we used diagnosis codes to identify solid organ or hematopoietic stem cell transplantation and HIV/AIDs versus patient registries. Consistent with the previously described algorithm, we required one inpatient or two outpatient diagnosis code for an immunocompromising condition (leukemia, lymphoma, congenital immunodeficiencies, asplenia/hyposplenia, HIV/AIDS, and organ transplant) in the year prior and any immunosuppressive medication (alkylating agents, antibiotics, antimetabolites, antimitotics, monoclonal antibodies, other, immune-modulating agents, TNF Alpha antagonist, and steroids) with an outpatient days supply or inpatient administration in the 90 days prior.
\*\*\*Medical history included underlying conditions and diagnoses in the year prior to the ARI episode, identified using international classification of diseases (ICD)-10 codes.

A higher proportion of those who tested negative for SARS-CoV-2 (compared to test-positive cases) had a Charlson Comorbidity Index of ≥4 (26.5% *vs* 22.0%; *P*<.001), were immunocompromised (37.3% *vs* 27.9%; *P*<.001), or were hospitalized in the past year (29.5% *vs* 23.2%; *P*<.001). These notable differences and other differences by case-control status are described in **Table 1**. A higher proportion of those who received the XBB vaccine (compared to those who did not) were ≥65 years of age (75.3% *vs* 50.2%; *P*<.001), were Black or African American (31.4% *vs* 25.9%; *P*<.001), had a Charlson Comorbidity Index of ≥4 (38.4% *vs* 24.8%; *P*<.001), or had previously received a BA.4/5-adapted bivalent vaccine (76.8% *vs* 22.9%; *P*<.001). These notable differences and other differences by XBB vaccination status are described in **Supplemental Table 2**.

Overall adjusted VE of BNT162b2 XBB vaccine was 43% (95% CI: 34–51%) against hospitalization (**Figure 2A**), 39% (33–45%) against ED/UC visits (**Figure 2B**), and 27% (16–37%) against outpatient visits (**Figure 2C**) compared to not receiving an XBB vaccine of any kind. Across all three outcomes VE declined after the likely XBB period during the JN.1 co-circulation and likely JN.1 periods, although CIs overlapped: 61% (44– 73%), 46% (32–58%), and 35% (20–48%), respectively, against hospitalization (**Figure 2A**); 50% (35–61%), 43% (33–52%), and 33% (22–43%), respectively, against ED/UC visits (**Figure 2B**); and 51% (27–67%), 29% (9–44%), and 24% (5–39%), respectively, against outpatient visits (**Figure 2C**).

**Figure 2.**
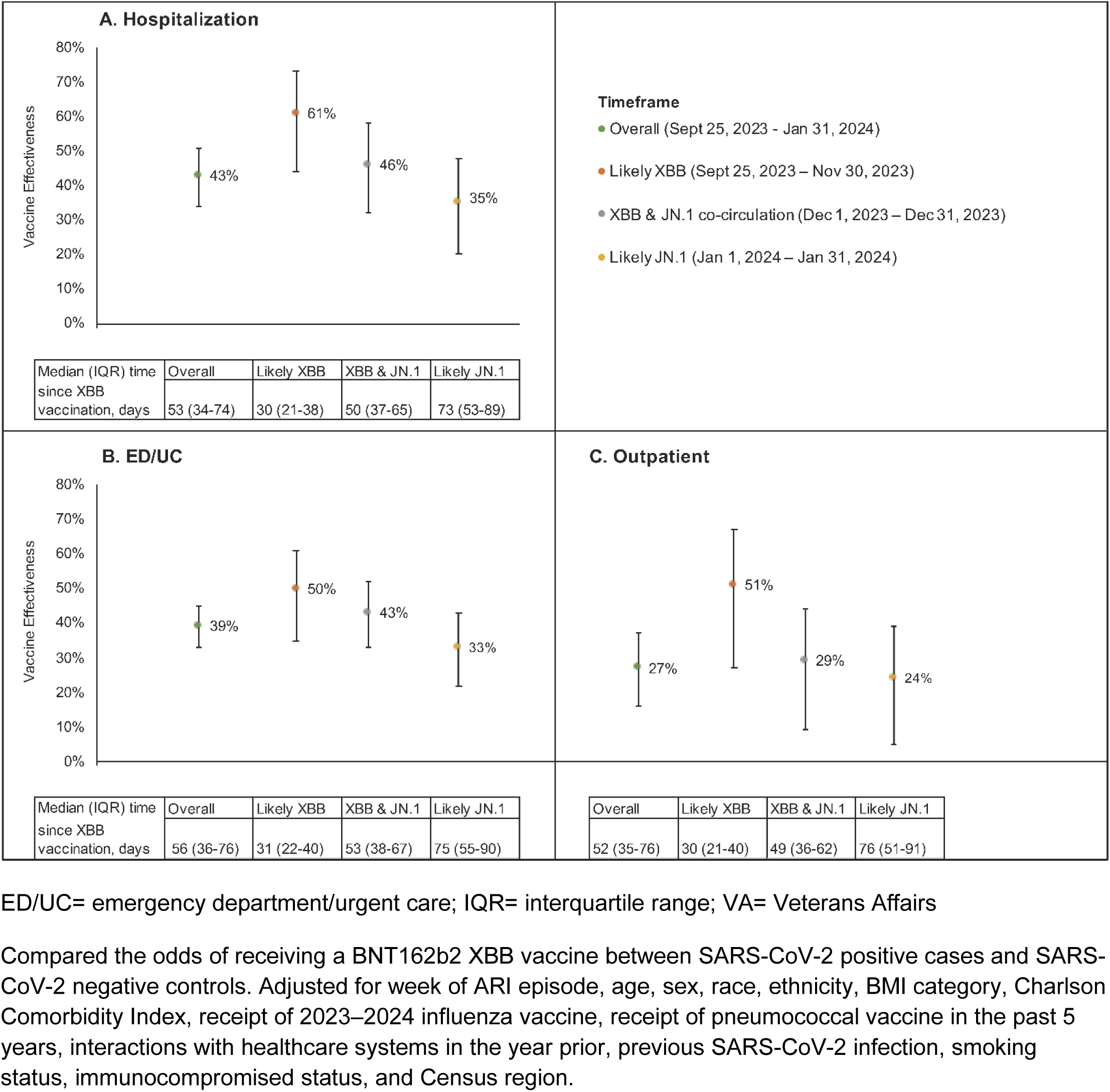
Adjusted vaccine effectiveness of the BNT162b2 XBB vaccine by time period for hospitalization (A), ED/UC visits (B), and outpatient visits (C)

Median time since receipt of BNT162b2 XBB vaccine was ≤76 days for all variant periods, but longer in the XBB and JN.1 co-circulation (51 days, IQR: 37–65) and likely JN.1 (75 days, IQR: 54–90) periods, than the likely XBB time period (30 days, IQR: 21–40). Thus, to help tease apart the impact of waning effectiveness and increasing prevalence of JN.1 over time, **Table 2** presents VE of the BNT162b2 XBB vaccine by both days since receipt of the vaccine and variant period. VE within 60 days of BNT162b2 XBB vaccination during the likely XBB period was 62% (95% CI 44–74%) against hospitalization, 52% (37–63%) against ED/UC visits, and 50% (25–66%) against outpatient visits. VE could not be calculated beyond 60 days of BNT162b2 XBB vaccination during the likely XBB period due to the small number of ARI episodes with a time since vaccination longer than 60 days during this time period. During the likely JN.1 period, VE within 60 days of vaccination was 32% (3–52%) against hospitalization, 41% (23–54%) against ED/UC visits, and 31% (1–52%) against outpatient visits, while VE 61–133 days since vaccination was 30% (16–41%) against ED/UC episodes and 20% (-4–38%) against outpatient visits.

**Table 2.** Adjusted vaccine effectiveness of the BNT162b2 XBB vaccine for hospitalization, ED/UC visits, and outpatient visits by variant period and time since vaccination.

| Outcome | Likely XBB<br>(Sep 25 – Nov 30, 2023) |  | Likely JN.1<br>(Jan 1 – Jan 31, 2024) |  |
| --- | --- | --- | --- | --- |
|  | VE (95% CI) by days since receipt of XBB dose |  |  |  |
|  | ≤ 60 | 61–133 | ≤ 60 | 61–133 |
| Hospitalization | 62 (44–74) | —* | 32 (3–52) | 37 (19–51) |
| ED/UC visit | 52 (37–63) | —* | 41 (23–54) | 30 (16–41) |
| Outpatient visit | 50 (25–66) | —* | 31 (1–52) | 20 (–4–38) |

CI= confidence interval; ED/UC= emergency department/urgent care; VE= vaccine effectiveness
Compared the odds of receiving a BNT162b2 XBB vaccine between SARS-CoV-2 positive cases and SARS-CoV-2 negative controls by days since vaccination. Adjusted for week of ARI episode, age, sex, race, ethnicity, BMI category, Charlson Comorbidity Index, receipt of 2023-2024 influenza vaccine, receipt of pneumococcal vaccine in the past 5 years, interactions with healthcare systems in the year prior, previous SARS-CoV-2 infection, smoking status, immunocompromised status, and Census region.
\*VE estimates based on 2 x 2 comparison tables with a cell size < 5 were not reported due to imprecision

**Supplemental tables 3, 4, 5, and 6** present VE results stratified by age group, immunocompromised status, obesity status, and smoking status, respectively. Although CIs overlapped across all stratified estimates, effectiveness estimates were 24–41% in those ≥65 years of age and 34–58% in those <65 years of age across all outcomes. VE was 33% against hospitalization and 34% against ED/UC visits in those who were immunocompromised and 49% and 42%, respectively, in those who were not. Finally, VE estimates were 34– 50% across all outcomes in those who were obese and 21–39% in those who were not, and VE was 51% against hospitalization in non-smokers and 38% among current or former smokers.

## DISCUSSION

In this test-negative case-control study conducted among a large national US Veteran population between September 25, 2023 and January 31, 2024, overall effectiveness of the BNT162b2 XBB vaccine compared to not receiving an XBB vaccine of any kind was 43% (95% CI: 34–51%) against COVID-19-associated hospitalization, 39% (33–45%) against ED/UC visits, and 27% (16–37%) against outpatient visits. These data add to a growing body of evidence that BNT162b2 XBB vaccine was effective at preventing a range of COVID- 19 outcomes during the 2023–2024 respiratory virus season.^15–19,29^ Notably, however, VE point estimates were lower for all three COVID-19 outcomes, including hospitalization, during the time period when COVID-19 was likely caused by JN.1 sub-lineages (24–35%) than when caused by XBB sub-lineages (50–61%), although CIs overlapped.

The observed reduction in VE during the likely JN.1 period did not appear to be driven by waning effectiveness over time for three reasons. First, all VE estimates in our study had a median time since receipt of a BNT162b2 XBB dose of ≤76 days. Thus, it seems unlikely that waning of protection would meaningfully impact our estimates given our relatively short follow-up period. Additionally, during the likely JN.1 period (when durability could be assessed), there appeared to be only very modest waning of effectiveness against JN.1 for less severe outcomes (i.e., ED/UC and outpatient visits) through a maximum of 133 days since BNT162b2 XBB vaccination. Finally, VE still appeared lower during the likely JN.1 period than the likely XBB period even when analyses were restricted to within 60 days of BNT162b2 XBB vaccination (32% [3–52%] and 62% [44–74%], respectively, against hospitalization; 41% [23–54%] and 52% [37–63%] against ED/UC visits; and 31% [1–52%] and 50% [25–66%] against outpatient visits).

These results suggest that XBB vaccines likely have reduced effectiveness against COVID-19 caused by JN.1 and its sub-lineages, which have now become predominant globally. Thus, like the last two years, a strain change for the upcoming 2024–2025 season also appears warranted to combat not only waning immunity over time, but reduced effectiveness stemming from continued SARS-CoV-2 evolution and vaccine strain mismatch. Reassuringly, annual updates to mRNA COVID-19 vaccines have restored protection against COVID-19 that has eroded over time due to a combination of waning of vaccine protection and the emergence of antigenically distinct strains.^9–13,15,17,29^ Thus, analogous to influenza, although prior versions of COVID-19 vaccines once provided high levels of protection, the combination of waning vaccine-induced immunity and continuous SARS-CoV-2 strain evolution eventually renders prior versions of vaccines less effective over time, even against severe clinical outcomes like hospitalization. This, in turn, warrants routine updates to COVID-19 vaccines, as with influenza, so long as SARS-CoV-2 continues to circulate and cause disease.

Other preliminary data also support our findings that XBB vaccines may have reduced effectiveness against JN.1. A recent test-negative design study conducted by CDC that evaluated the effectiveness of XBB vaccines against symptomatic COVID-19 detected in the pharmacy setting between September 21, 2023 and January 14, 2024 suggested that VE against JN.1 sub-lineages may be lower than XBB sub-lineages (49% [19–68%] and 60% [35–75%], respectively), although variant-specific CIs overlapped in this analysis as well.^17^ In addition, when looking at the totality of XBB VE data published to date globally, there appears to be a general decline in VE over time—corresponding to increases in the prevalence of JN.1 over the same time period. For example, early reports showed >70% effectiveness for XBB vaccines against COVID-19 hospitalization during time periods when JN.1 was not circulating or was present only at low levels.^16,19^ However, subsequent reports with longer study periods that included more JN.1 co-circulation have shown sequentially lower VE. A report from Kaiser Permanente Southern California that included data through mid-December showed a slightly lower overall VE of roughly 60% against COVID-19 hospitalization,^15^ and a recent CDC report that included data through the end of January 2024 showed roughly 40–50% VE against COVID-19 hospitalization.^29^ This latter CDC estimate^29^ is similar to the 40% overall VE against hospitalization we observed in this study which also included data through the end of January 2024. Finally, these VE data are consistent with data showing that JN.1 is phylogenetically and antigenically distant from XBB sub-lineages,^21,22^ and that neutralization activity of XBB vaccines is lower against JN.1 compared to compared to XBB strains.^23^

It is possible that the lower overall VE observed in our study may also be explained by the national VA population, which is generally older, predominantly male, and with a high prevalence of multiple comorbid conditions.^30^ Reductions in vaccine immunogenicity and effectiveness with increasing age have been demonstrated previously,^31^ and we observed slightly lower effectiveness among individuals ≥65 years of age in our study. Male sex and comorbidities are also well-established risk factors for severe COVID-19.^32,33^ Further, our study differed from previous VE studies by including both NAAT and RAT to identify SARS-CoV- 2 rather than NAAT alone. Sensitivity of RAT is lower than molecular testing, particularly as population immunity against SARS-CoV-2 has increased over time.^34,35^ Although the inclusion of RAT allowed a more robust outpatient sample to be included in the study and represents real world testing behaviors, an under-detection of cases (due to potential reduced RAT sensitivity) could bias VE estimates towards the null presuming non-differential misclassification across vaccination status. Despite these study population and design differences that could partially explain the slightly lower overall VE observed in our study compared with previous reports, these differences would not explain the marked differences in point estimates for VE against XBB-related and JN.1-related COVID-19 we saw in our study population across multiple outcomes. Finally, ours and other recent XBB VE estimates, were derived from study populations where nearly all participants had some level of pre-existing COVID-19 immunity stemming from prior vaccination and infection,^36^ which may result in lower apparent effectiveness compared to the initial COVID-19 vaccine rollout when levels of baseline immunity were lower.^37^

There are several important limitations to consider when interpreting these data. The test-negative case-control study, while considered a reliable design for evaluating real-world VE due to its ability to mitigate both healthcare-seeking and testing behavior, is still susceptible to selection bias.^38–40^ Namely, although we employed statistical adjustments to control for potentially confounding patient clinical and sociodemographic characteristics, there may still have been residual confounding by unmeasured factors. We accounted for SARS-CoV-2 activity and circulating strain by adjusting for calendar week and performing analyses that were stratified by variant time period, however, this may not have fully accounted for increased SARS-CoV-2 activity in December 2023 and January 2024.^41^ It is also important to note that our VE estimates against XBB or JN.1 sub-lineages relied on time periods rather than sequencing data. However, our variant time period estimates are likely still a valid approximation of strain-specific VE because XBB and JN.1 sub-lineages accounted for >90% and >80% of sequenced SARS-CoV-2 strains in the United States during our likely XBB and likely JN.1 periods, respectively.^26^ Another limitation is that we were unable to evaluate VE beyond roughly 4 months since receipt of a BNT162b2 XBB dose given our study period. Thus, additional analyses of longer-term durability are needed. Similarly, our analysis, like others before it, was unable to evaluate durability of VE against XBB sub-lineages given they were overtaken by JN.1 fewer than 90 days after vaccine rollout.^15–19,29^

We were also unable to assess whether VE estimates differed between those tested by NAAT versus RAT, as the type of laboratory test is not uniformly captured in the VA database. Additionally, it is possible that some of the SARS-CoV-2-positive ARI episodes were incidental infections (i.e., sought care “with COVID-19” rather than “for COVID-19"), which could also lead to underestimation of VE. The accuracy of XBB vaccination status depended on the completeness and reliability of the vaccination records within the VA system. However, VA data capture most vaccines given outside of the VA, and any rare instances of uncaptured COVID-19 vaccine administrations would likely result in underestimation of VE. Finally, our findings may not be generalizable to the general US population or globally, due to the unique characteristics of the VA population that include a higher proportion of patients that are older, male, and have underlying chronic medical conditions. Veterans may also exhibit unique healthcare-seeking behavior given their access to the VA Healthcare System.

## CONCLUSIONS

BNT162b2 XBB vaccine was effective at preventing a range of COVID-19 outcomes during the 2023–2024 respiratory virus season. However, VE appeared lower during a period when most COVID-19 cases were caused by JN.1 even after accounting for time since receipt of a BNT162b2 XBB dose. These data underscore the importance of strain match to maximize the public health impact of COVID-19 vaccination.

## Data Availability

The study data may be made available upon reasonable request and approval by the Department of Veterans Affairs.

## Author Contribution

Conception and design of the study: ARC, HJA, VVL, LP, EJZ, JMM

Data generation: ARC, HJA, VVL

Analysis and/or interpretation of the data: All authors

Preparation or critical revision of the manuscript: All authors

## Ethics Approval

This study was determined to be exempt by the VA Providence Healthcare System (VAPHS) Institutional Review Board (IRB) and approved by the VAPHS Research and Development Committee. As this was a retrospective study of existing health records and exempt from IRB review, informed consent requirements are not applicable.

## Other Acknowledgements

Views expressed are those of the authors and do not necessarily reflect the position or policy of the United States Department of Veterans Affairs.

The research study would not have been possible without the health information from patients under the care of the Veterans Health Administration. We express our gratitude to the VA patients for their invaluable contributions to medical and scientific progress.

