## Supplemental Materials for "Effectiveness of BNT162b2 XBB vaccine in the US Veterans Affairs Healthcare System"

### Supplemental Information for Effectiveness of BNT162b2 XBB vaccine in the US Veterans Affairs Healthcare System

**Supplemental Table 1.** Acute respiratory infection diagnosis codes (ICD-10)

| ICD-10 Code | Diagnosis |
|---|---|
| A22.1 | Pulmonary anthrax |
| A37.00 | Whooping cough due to *Bordetella pertussis* without pneumonia |
| A37.01 | Whooping cough due to *Bordetella pertussis* with pneumonia |
| A37.10 | Whooping cough due to *Bordetella parapertussis* without pneumonia |
| A37.11 | Whooping cough due to *Bordetella parapertussis* with pneumonia |
| A37.80 | Whooping cough due to other Bordetella species without pneumonia |
| A37.81 | Whooping cough due to other Bordetella species with pneumonia |
| A37.90 | Whooping cough, unspecified species without pneumonia |
| A37.91 | Whooping cough, unspecified species with pneumonia |
| A48.1 | Legionnaires' disease |
| B25.0 | Cytomegaloviral pneumonitis |
| B34.2 | Coronavirus infection, unspecified |
| B44.0 | Invasive pulmonary aspergillosis |
| B77.81 | Ascariasis pneumonia |
| B97.29 | Other coronavirus as the cause of diseases classified elsewhere |
| J00* | Acute nasopharyngitis [common cold] |
| J01* | Acute sinusitis |
| J02* | Acute pharyngitis |
| J03* | Acute tonsillitis |
| J04* | Acute laryngitis and tracheitis |
| J05* | Acute obstructive laryngitis [croup] and epiglottitis |
| J06* | Acute upper respiratory infections of multiple and unspecified sites |
| J09.X1 | Influenza due to novel influenza a virus with pneumonia |
| J09.X2 | Influenza due to identified novel influenza A virus with other respiratory manifestations |
| J09.X3 | Influenza due to identified novel influenza A virus with gastrointestinal manifestations |
| J09.X9 | Influenza due to identified novel influenza A virus with other manifestations |
| J10.00 | Influenza due to other identified influenza virus with pneumonia |
| J10.01 | Influenza due to other identified influenza virus with the same other identified influenza virus pneumonia |
| J10.08 | Influenza due to other identified influenza virus with other specified pneumonia |
| J10.1 | Influenza due to other identified influenza virus with other respiratory manifestations |
| J10.2 | Influenza due to other identified influenza virus with gastrointestinal manifestations |
| J10.8* | Influenza due to other identified influenza virus with other manifestations |
| J10.81 | Influenza due to other identified influenza virus with encephalopathy |
| J10.82 | Influenza due to other identified influenza virus with myocarditis |
| J10.83 | Influenza due to other identified influenza virus with otitis media |
| J10.89 | Influenza due to other identified influenza virus with other manifestations |
| J11.00 | Influenza due to unidentified influenza virus with pneumonia |
| J11.08 | Influenza due to unidentified influenza virus with specified pneumonia |
| J11.1 | Influenza due to unidentified influenza virus with other respiratory manifestations |
| J11.2 | Influenza due to unidentified influenza virus with gastrointestinal manifestations |
| J11.81 | Influenza due to unidentified influenza virus with encephalopathy |
| J11.82 | Influenza due to unidentified influenza virus with myocarditis |
| J11.83 | Influenza due to unidentified influenza virus with otitis media |
| J11.89 | Influenza due to unidentified influenza virus with other manifestations |
| J12.0 | Adenoviral pneumonia |
| J12.1 | Respiratory syncytial virus pneumonia |
| J12.2 | Parainfluenza virus pneumonia |
| J12.3 | Human metapneumovirus pneumonia |
| J12.81 | Pneumonia due to SARS-associated coronavirus |
| J12.82 | Pneumonia due to coronavirus disease 2019 |
| J12.89 | Other viral pneumonia |
| J12.9 | Viral pneumonia, unspecified |
| J13 | Pneumonia due to *Streptococcus pneumoniae* |
| J14 | Pneumonia due to *Hemophilus influenzae* |

| ICD-10 Code | Diagnosis |
|---|---|
| J15.0 | Pneumonia due to *Klebsiella pneumoniae* |
| J15.1 | Pneumonia due to Pseudomonas |
| J15.20 | Pneumonia due to Staphylococcus, unspecified |
| J15.211 | Pneumonia due to methicillin susceptible *Staphylococcus aureus* |
| J15.212 | Pneumonia due to methicillin resistant *Staphylococcus aureus* |
| J.15.29 | Pneumonia due to other Staphylococcus |
| J15.3 | Pneumonia due to Streptococcus, group b |
| J15.4 | Pneumonia due to other Streptococci |
| J15.5 | Pneumonia due to *Escherichia coli* |
| J15.6 | Pneumonia due to other aerobic gram-negative bacteria |
| J15.7 | Pneumonia due to *Mycoplasma pneumoniae* |
| J15.8 | Pneumonia due to other specified bacteria |
| J15.9 | Unspecified bacterial pneumonia |
| J16.0 | Chlamydial pneumonia |
| J16.8 | Pneumonia due to other specified infectious organisms |
| J17 | Pneumonia in diseases classified elsewhere |
| J18.0 | Bronchopneumonia, unspecified organism |
| J18.1 | Lobar pneumonia, unspecified organism |
| J18.2 | Hypostatic pneumonia, unspecified organism |
| J18.8 | Other pneumonia, unspecified organism |
| J18.9 | Pneumonia, unspecified organism |
| J20.0 | Acute bronchitis due to *Mycoplasma pneumoniae* |
| J20.1 | Acute bronchitis due to *Hemophilus influenzae* |
| J20.2 | Acute bronchitis due to Streptococcus |
| J20.3 | Acute bronchitis due to coxsackievirus |
| J20.4 | Acute bronchitis due to parainfluenza virus |
| J20.5 | Acute bronchitis due to respiratory syncytial virus |
| J20.6 | Acute bronchitis due to rhinovirus |
| J20.7 | Acute bronchitis due to echovirus |
| J20.8 | Acute bronchitis due to other specified organisms |
| J20.9 | Acute bronchitis, unspecified |
| J21.* | Acute bronchiolitis |
| J21.0 | Acute bronchiolitis due to respiratory syncytial virus |
| J21.1 | Acute bronchiolitis due to human metapneumovirus |
| J21.8 | Acute bronchiolitis due to other specified organisms |
| J21.9 | Acute bronchiolitis, unspecified |
| J22 | Unspecified acute lower respiratory infection |
| J80 | Acute respiratory distress syndrome |
| J96.00 | Acute respiratory failure unspecified whether with hypoxia or hypercapnia |
| J96.01 | Acute respiratory failure with hypoxia |
| J96.02 | Acute respiratory failure with hypercapnia |
| J96.10 | Chronic respiratory failure, unspecified with hypoxia or hypercapnia |
| J96.11 | Chronic respiratory failure with hypoxia |
| J96.12 | Chronic respiratory failure with hypercapnia |
| J96.20 | Acute and chr resp failure, unspecified with hypoxia or hypercapnia |
| J96.21 | Acute and chronic respiratory failure with hypoxia |
| J96.22 | Acute and chronic respiratory failure with hypercapnia |
| J96.90 | Respiratory failure, unspecified, unspecified with hypoxia or hypercapnia |
| J96.91 | Respiratory failure, unspecified with hypoxia |
| J96.92 | Respiratory failure, unspecified with hypercapnia |
| M35.81 | Multisystem inflammatory syndrome |
| R04.2 | Hemoptysis |
| R05 | Cough |
| R05.1 | Acute cough |
| R05.2 | Subacute cough |
| R05.3 | Chronic cough |
| R05.4 | Cough syncope |

| ICD-10 Code | Diagnosis |
|---|---|
| R05.8 | Other specified cough |
| R05.9 | Cough, unspecified |
| R06.00 | Dyspnea/abnormalities of breathing unspecified |
| R06.02 | Shortness of breath |
| R06.03 | Acute respiratory distress |
| R06.09 | Other forms of dyspnea |
| R06.1 | Stridor |
| R06.82 | Tachypnea, not elsewhere classified |
| R06.89 | Other abnormalities of breathing |
| R07.1 | Chest pain on breathing |
| R09.0* | Asphyxia and hypoxemia |
| R09.01 | Asphyxia |
| R09.02 | Hypoxemia |
| R09.1 | Pleurisy |
| R09.2 | Respiratory arrest |
| R50.9 | Fever, unspecified |
| U04* | SARS (WHO 2019) |
| u04.9 | SARS, unspecified (WHO 2019) |
| U07.1 | COVID-19 |
| U07.2 | COVID-19, virus not identified (clinically diagnosed) |

**Supplemental Table 2.** Demographics and clinical characteristics of acute respiratory infection episodes (hospitalization, ED/UC visits, outpatient visits) with SARS-CoV-2 testing by vaccination status

| Variable | Total (n=113,174) | Received BNT162b2 XBB vaccine (n=7,324) | No XBB vaccine of any kind (n=105,850) | *P*-value |
|---|---|---|---|---|
| **COVID vaccine status** ≥1 dose of BA.4/5-adapted bivalent vaccine | 29,907 (26.4) | 5,626 (76.8) | 24,281 (22.9) | <0.001 |
| ≥3 doses of original wild-type mRNA vaccine but no variant-adapted vaccines | 29,463 (26.0) | 1,262 (17.2) | 28,201 (26.6) | <0.001 |
| ≥2 doses of original wild-type mRNA vaccine but no variant-adapted vaccines | 51,546 (45.6) | 1,499 (20.5) | 50,047 (47.3) | <0.001 |
| Unvaccinated | 24,747 (21.9) | 106 (1.4) | 24,641 (23.3) | <0.001 |
| Time since last non-XBB adapted vaccine, median days (IQR) | 670 (413–820) | 433 (385–477) | 691 (421–840) | <0.001 |
| **Age group** 18–64 years | 54,563 (48.2) | 1,811 (24.7) | 52,752 (49.8) | <0.001 |
| 65–74 years | 27,821 (24.6) | 2,416 (33) | 25,405 (24) | |
| ≥75 years | 30,790 (27.2) | 3,097 (42.3) | 27,693 (26.2) | |
| **Sex** Male | 98,172 (86.7) | 6,644 (90.7) | 91,528 (86.5) | <0.001 |
| Female | 15,002 (13.3) | 680 (9.3) | 14,322 (13.5) | |
| **Body mass index category** Underweight (<18.5 kg/m$^2$) | 1,399 (1.2) | 64 (0.9) | 1,335 (1.3) | <0.001 |
| Healthy weight (18.5–24.9 kg/m$^2$) | 34,083 (30.1) | 2,252 (30.7) | 31,831 (30.1) | |
| Overweight (25–29.9 kg/m$^2$) | 22,780 (20.2) | 1,571 (21.5) | 21,209 (20) | |

| Variable | Total (n=113,174) | Received BNT162b2 XBB vaccine (n=7,324) | No XBB vaccine of any kind (n=105,850) | *P*-value |
|---|---|---|---|---|
| Obese (≥30 kg/m$^2$) | 54,398 (48.3) | 3,429 (46.8) | 50,969 (48.2) | |
| **Region** | | | | <0.001 |
| Midwest | 22,167 (19.6) | 1,830 (25) | 20,337 (19.2) | |
| Northeast | 15,063 (13.3) | 1,050 (14.3) | 14,013 (13.2) | |
| West | 21,933 (19.4) | 1,458 (19.9) | 20,475 (19.3) | |
| South | 54,011 (47.7) | 2,986 (40.8) | 51,025 (48.2) | |
| **Race** | | | | <0.001 |
| Black or African American | 29,699 (26.2) | 2,301 (31.4) | 27,398 (25.9) | |
| White | 71,345 (63.0) | 4,337 (59.2) | 67,008 (63.3) | |
| Other race | 12,130 (10.7) | 686 (9.4) | 11,444 (10.8) | |
| **Ethnicity** | | | | <0.001 |
| Hispanic or Latino | 10,527 (9.3) | 472 (6.4) | 10,055 (9.5) | |
| Not Hispanic or Latino | 102,647 (90.7) | 6,852 (93.6) | 95,795 (90.5) | |
| **Smoking** | | | | <0.001 |
| Current or former | 58,062 (51.3) | 4,122 (56.3) | 53,940 (51) | |
| **Area deprivation index (ADI) Quintile** | | | | <0.001 |
| 1 (Least Deprived) | 22,140 (19.6) | 1,845 (25.2) | 20,295 (19.2) | |
| 2 | 22,134 (19.6) | 1,396 (19.1) | 20,738 (19.6) | |
| 3 | 22,143 (19.6) | 1,357 (18.5) | 20,786 (19.6) | |
| 4 | 22,126 (19.6) | 1,243 (17) | 20,883 (19.7) | |
| 5 (Most Deprived) | 22,136 (19.6) | 1,415 (19.3) | 20,721 (19.6) | |
| **VA Frailty index (VA-FI)*** | | | | <0.001 |
| Non-frail (VA-FI ≤0.1) | 40,182 (35.5) | 1,435 (19.6) | 38,747 (36.6) | |
| Pre-frail (VA-FI >0.1-0.2) | 27,422 (24.2) | 1,721 (23.5) | 25,701 (24.3) | |
| Mildly frail (VA-FI >0.2-0.3) | 18,818 (16.6) | 1,597 (21.8) | 17,221 (16.3) | |

| Variable | Total (n=113,174) | Received BNT162b2 XBB vaccine (n=7,324) | No XBB vaccine of any kind (n=105,850) | *P*-value |
|---|---|---|---|---|
| Moderately frail (VA-FI >0.3-0.4) | 12,559 (11.1) | 1,129 (15.4) | 11,430 (10.8) | |
| Severely frail (VA-FI ≥0.5) | 14,193 (12.5) | 1,442 (19.7) | 12,751 (12) | |
| **Healthcare exposures, 1 year prior** Hospital admission | 32,067 (28.3) | 2,599 (35.5) | 29,468 (27.8) | <0.001 |
| Nursing home admission | 3,924 (3.5) | 325 (4.4) | 3,599 (3.4) | <0.001 |
| Intensive care unit admission | 8,825 (7.8) | 685 (9.4) | 8,140 (7.7) | <0.001 |
| Emergency department visit | 69,978 (61.8) | 4,797 (65.5) | 65,181 (61.6) | <0.001 |
| Primary care visit | 106,691 (94.3) | 7,165 (97.8) | 99,526 (94) | <0.001 |
| **Charlson Comorbidity Index** 0 | 38,598 (34.1) | 1,370 (18.7) | 37,228 (35.2) | <0.001 |
| 1 | 21,698 (19.2) | 1,268 (17.3) | 20,430 (19.3) | |
| 2 | 12,962 (11.5) | 952 (13) | 12,010 (11.3) | |
| 3 | 10,833 (9.6) | 919 (12.5) | 9,914 (9.4) | |
| ≥ 4 | 29,083 (25.7) | 2,815 (38.4) | 26,268 (24.8) | |
| Immunocompromised** | 40,309 (35.6) | 2,961 (40.4) | 37,348 (35.3) | <0.001 |
| **Medical History***** Acute cerebrovascular disease | 5,372 (4.7) | 467 (6.4) | 4,905 (4.6) | <0.001 |
| Acute myocardial infarction | 3,842 (3.4) | 301 (4.1) | 3,541 (3.3) | <0.001 |
| Alcohol and substance related disorders | 28,011 (24.8) | 1,672 (22.8) | 26,339 (24.9) | <0.001 |
| Any cancer or malignancy | 42,527 (37.6) | 3,695 (50.5) | 38,832 (36.7) | <0.001 |
| Aortic and peripheral arterial embolism or thrombosis | 440 (0.4) | 47 (0.6) | 393 (0.4) | <0.001 |

| Variable | Total (n=113,174) | Received BNT162b2 XBB vaccine (n=7,324) | No XBB vaccine of any kind (n=105,850) | *P*-value |
|---|---|---|---|---|
| Asthma | 10,024 (8.9) | 759 (10.4) | 9,265 (8.8) | <0.001 |
| Benign prostatic hyperplasia | 23,037 (20.4) | 2,201 (30.1) | 20,836 (19.7) | <0.001 |
| Cardiac dysrhythmias | 27,478 (24.3) | 2,304 (31.5) | 25,174 (23.8) | <0.001 |
| Chronic kidney disease | 13,161 (11.6) | 1,313 (17.9) | 11,848 (11.2) | <0.001 |
| Chronic obstructive pulmonary disease and bronchiectasis | 26,259 (23.2) | 2,247 (30.7) | 24,012 (22.7) | <0.001 |
| Congestive heart failure | 17,429 (15.4) | 1,589 (21.7) | 15,840 (15) | <0.001 |
| Coronary atherosclerosis and other heart disease | 25,404 (22.4) | 2,225 (30.4) | 23,179 (21.9) | <0.001 |
| Delirium, dementia, and other cognitive disorders | 9,559 (8.4) | 882 (12) | 8,677 (8.2) | <0.001 |
| Diabetes with or without chronic complications | 47,147 (41.7) | 3,885 (53) | 43,262 (40.9) | <0.001 |
| Epilepsy | 3,142 (2.8) | 228 (3.1) | 2,914 (2.8) | 0.070 |
| Human immunodeficiency virus (HIV) infection | 1,271 (1.1) | 140 (1.9) | 1,131 (1.1) | <0.001 |
| Hypertension | 66,411 (58.7) | 5,339 (72.9) | 61,072 (57.7) | <0.001 |
| Influenza | 1,413 (1.2) | 65 (0.9) | 1,348 (1.3) | 0.004 |
| Liver diseases | 10,877 (9.6) | 827 (11.3) | 10,050 (9.5) | <0.001 |
| Mental health conditions | 57,479 (50.8) | 3,571 (48.8) | 53,908 (50.9) | <0.001 |
| Osteoarthritis | 23,617 (20.9) | 2,105 (28.7) | 21,512 (20.3) | <0.001 |
| Peripheral and visceral atherosclerosis | 9,699 (8.6) | 926 (12.6) | 8,773 (8.3) | <0.001 |
| Pneumonia | 9,596 (8.5) | 774 (10.6) | 8,822 (8.3) | <0.001 |
| Pulmonary heart disease | 7,598 (6.7) | 674 (9.2) | 6,924 (6.5) | <0.001 |
| Rheumatoid arthritis | 2,273 (2.0) | 220 (3) | 2,053 (1.9) | <0.001 |
| Septicemia | 4,805 (4.2) | 390 (5.3) | 4,415 (4.2) | <0.001 |

| Variable | Total (n=113,174) | Received BNT162b2 XBB vaccine (n=7,324) | No XBB vaccine of any kind (n=105,850) | *P*-value |
|---|---|---|---|---|
| Thyroid disorder | 14,098 (12.5) | 1,113 (15.2) | 12,985 (12.3) | <0.001 |
| Tuberculosis | 237 (0.2) | 21 (0.3) | 216 (0.2) | 0.134 |
| **Week of infection** | | | | <0.001 |
| Sep 25–Sep 30,2023 | 5,503 (4.9) | < 5 (<0.1) | 5,503 (5.2) | |
| Oct 01–Oct 07, 2023 | 4,834 (4.3) | 6 (0.1) | 4,828 (4.6) | |
| Oct 08–Oct 14, 2023 | 4,358 (3.9) | 7 (0.1) | 4,351 (4.1) | |
| Oct 15–Oct 21, 2023 | 4,707 (4.2) | 31 (0.4) | 4,676 (4.4) | |
| Oct 22–Oct 28, 2023 | 4,624 (4.1) | 74 (1) | 4,550 (4.3) | |
| Oct 29–Nov 04, 2023 | 4,453 (3.9) | 121 (1.7) | 4,332 (4.1) | |
| Nov 05–Nov 11, 2023 | 4,741 (4.2) | 201 (2.7) | 4,540 (4.3) | |
| Nov 12–Nov 18, 2023 | 5,622 (5.0) | 291 (4) | 5,331 (5) | |
| Nov 19–Nov 25, 2023 | 5,155 (4.6) | 320 (4.4) | 4,835 (4.6) | |
| Nov 26–Dec 02, 2023 | 6,746 (6.0) | 474 (6.5) | 6,272 (5.9) | |
| Dec 03–Dec 09, 2023 | 6,855 (6.1) | 524 (7.2) | 6,331 (6) | |
| Dec 10–Dec 16, 2023 | 6,959 (6.1) | 568 (7.8) | 6,391 (6) | |
| Dec 17–Dec 23, 2023 | 8,160 (7.2) | 680 (9.3) | 7,480 (7.1) | |
| Dec 24–Dec 30, 2023 | 8,366 (7.4) | 792 (10.8) | 7,574 (7.2) | |
| Dec 31, 2023–Jan 06, 2024 | 8,814 (7.8) | 871 (11.9) | 7,943 (7.5) | |
| Jan 07–Jan 13, 2024 | 7,624 (6.7) | 759 (10.4) | 6,865 (6.5) | |
| Jan 14–Jan 20, 2024 | 5,663 (5.0) | 579 (7.9) | 5,084 (4.8) | |
| Jan 21–Jan 27, 2024 | 6,316 (5.6) | 623 (8.5) | 5,693 (5.4) | |
| Jan 28–Jan 31, 2024 | 3,674 (3.2) | 403 (5.5) | 3,271 (3.1) | |
| Prior COVID-19 infection | 31,195 (27.6) | 1,936 (26.4) | 29,259 (27.6) | 0.025 |
| Virtual visit (outpatient only) | 2,773 (10.3) | 204 (11.1) | 2,569 (10.2) | 0.221 |
| ICU admission (hospitalized only) | 4,955 (20.5) | 370 (20.2) | 4,585 (20.5) | 0.726 |
| Current influenza vaccine | 39,077 (34.5) | 6,779 (92.6) | 32,298 (30.5) | <0.001 |

| Variable | Total (n=113,174) | Received BNT162b2 XBB vaccine (n=7,324) | No XBB vaccine of any kind (n=105,850) | *P*-value |
|---|---|---|---|---|
| Pneumococcal vaccine in last 5 years | 41,032 (36.3) | 3,801 (51.9) | 37,231 (35.2) | <0.001 |

ARI= acute respiratory infection; ED/UC= emergency department/urgent care; VA= Veterans Affairs

Data are n (%) unless otherwise specified. Chi-square or Fisher's Exact tests were used to compare differences in proportions between the groups. For continuous variables, comparisons were performed using the Wilcoxon Rank Sum test or a Student's t-test, depending on the distribution of the data for the given variable.

All ARI encounters within a 30-day window were considered a single ARI episode. If multiple encounter types occurred during the 30-day window, the highest level of care was used (hospitalization > ED/UC > outpatient).

*VA Frailty index was categorized as non-frail (VA-FI ≤ 0.1), prefrail (>0.1–0.2), mildly frail (>0.2–0.3), moderately frail (>0.3–0.4), and severely frail (>0.4)

**Immunocompromised status was based on immunocompromising conditions in the year prior and immunosuppressive medications in the 90 days prior to the ARI episode based on a slightly modified algorithm that has been previously described. (Tartof SY, et al. Lancet Reg Health Am. 2022:9:100198.) Unlike the previously described algorithm, we used diagnosis codes to identify solid organ or hematopoietic stem cell transplantation and HIV/AIDs versus patient registries. Consistent with the previously described algorithm, we required one inpatient or two outpatient diagnosis code for an immunocompromising condition (leukemia, lymphoma, congenital immunodeficiencies, asplenia/hyposplenia, HIV/AIDS, and organ transplant) in the year prior and any immunosuppressive medication (alkylating agents, antibiotics, antimetbolites, antimitotics, monoclonal antibodies, other, immune-modulating agents, TNF Alpha antagonist, and steroids) with an outpatient days supply or inpatient administration in the 90 days prior.

***Medical history included underlying conditions and diagnoses in the year prior to the ARI episode, identified using international classification of diseases (ICD)-10 codes.

**Supplemental Table 3.** Adjusted vaccine effectiveness of the BNT162b2 XBB vaccine for hospitalization, ED/UC visits, and outpatient visits by age group

| Outcome | Age ≥65 years (n = 58,611) | | Age <65 years (n = 54,563) | |
|---|---|---|---|---|
| | VE (95% CI) | Median (IQR) days since XBB vaccine | VE (95% CI) | Median (IQR) days since XBB vaccine |
| Hospitalization | 41 (32–50) | 54 (33–74) | 58 (33–73) | 50 (34–67) |
| ED/UC visit | 35 (27–43) | 56 (36–77) | 48 (37–57) | 54 (35–74) |
| Outpatient visit | 24 (9–36) | 53 (35–76) | 34 (14–50) | 51 (33–75) |

CI= confidence interval; ED/UC= emergency department/urgent care; IQR= interquartile range; VA= Veterans Affairs; VE = vaccine effectiveness

Compared the odds of receiving a BNT162b2 XBB vaccine between SARS-CoV-2 positive cases and SARS-CoV-2 negative controls. Adjusted for week of ARI episode, age, sex, race, ethnicity, BMI category, Charlson Comorbidity Index, receipt of 2023–2024 influenza vaccine, receipt of pneumococcal vaccine in the past 5 years, interactions with healthcare systems in the year prior, previous SARS-CoV-2 infection, smoking status, immunocompromised status, and Census region.

**Supplemental Table 4.** Adjusted vaccine effectiveness of the BNT162b2 XBB vaccine for hospitalization, ED/UC visits, and outpatient visits by immunocompromised status

| Outcome | Immunocompromised (*n* = 40,309) | | Not Immunocompromised (*n* = 72,865) | |
|---|---|---|---|---|
| | VE (95% CI) | Median (IQR) days since XBB vaccine | VE (95% CI) | Median (IQR) days since XBB vaccine |
| Hospitalization | 33 (16–47) | 52 (33–73) | 49 (38–58) | 54 (34–74) |
| ED/UC visit | 34 (22–45) | 55 (35–74) | 42 (34–49) | 56 (36–77) |
| Outpatient visit | 40 (19–55) | 54 (35–77) | 22 (8–34) | 52 (34–75) |

CI= confidence interval; ED/UC= emergency department/urgent care; IQR= interquartile range; VA= Veterans Affairs; VE = vaccine effectiveness

Compared the odds of receiving a BNT162b2 XBB vaccine between SARS-CoV-2 positive cases and SARS-CoV-2 negative controls. Adjusted for week of ARI episode, age, sex, race, ethnicity, BMI category, Charlson Comorbidity Index, receipt of 2023–2024 influenza vaccine, receipt of pneumococcal vaccine in the past 5 years, interactions with healthcare systems in the year prior, previous SARS-CoV-2 infection, smoking status, immunocompromised status, and Census region.

.

**Supplemental Table 5.** Adjusted vaccine effectiveness of the BNT162b2 XBB vaccine for hospitalization, ED/UC visits, and outpatient visits by obesity classification

| Outcome | Obese (*n* = 54,398) | | Non-obese (*n* = 58,262) | |
|---|---|---|---|---|
| | VE (95% CI) | Median (IQR) days since XBB vaccine | VE (95% CI) | Median (IQR) days since XBB vaccine |
| Hospitalization | 50 (36–61) | 52 (33–74) | 39 (27–49) | 53 (34–73) |
| ED/UC visit | 44 (35–52) | 56 (36–76) | 35 (25–43) | 56 (36–76) |
| Outpatient visit | 34 (19–47) | 54 (35–76) | 21 (3–35) | 52 (34–76) |

CI= confidence interval; ED/UC= emergency department/urgent care; IQR= interquartile range; VA= Veterans Affairs; VE = vaccine effectiveness

Compared the odds of receiving a BNT162b2 XBB vaccine between SARS-CoV-2 positive cases and SARS-CoV-2 negative controls. Adjusted for week of ARI episode, age, sex, race, ethnicity, BMI category, Charlson Comorbidity Index, receipt of 2023–2024 influenza vaccine, receipt of pneumococcal vaccine in the past 5 years, interactions with healthcare systems in the year prior, previous SARS-CoV-2 infection, smoking status, immunocompromised status, and Census region.

**Supplemental Table 6.** Adjusted vaccine effectiveness of the BNT162b2 XBB vaccine for hospitalization, ED/UC visits, and outpatient visits by smoking status

| Outcome | Current or former smoker (*n* = 58,062) | | Non-smoker (*n* = 55,112) | |
|---|---|---|---|---|
| | VE (95% CI) | Median (IQR) days since XBB vaccine | VE (95% CI) | Median (IQR) days since XBB vaccine |
| Hospitalization | 38 (26–48) | 53 (34–73) | 51 (38–62) | 53 (33–75) |
| ED/UC visit | 43 (35–51) | 55 (35–76) | 34 (24–43) | 56 (37–77) |
| Outpatient visit | 32 (17–45) | 52 (33–74) | 22 (4–36) | 54 (36–77) |

CI= confidence interval; ED/UC= emergency department/urgent care; IQR= interquartile range; VA= Veterans Affairs; VE = vaccine effectiveness

Compared the odds of receiving a BNT162b2 XBB vaccine between SARS-CoV-2 positive cases and SARS-CoV-2 negative controls. Adjusted for week of ARI episode, age, sex, race, ethnicity, BMI category, Charlson Comorbidity Index, receipt of 2023–2024 influenza vaccine, receipt of pneumococcal vaccine in the past 5 years, interactions with healthcare systems in the year prior, previous SARS-CoV-2 infection, smoking status, immunocompromised status, and Census region.
